# Gene-environment interaction analysis incorporating sex, cardiometabolic diseases, and multiple deprivation index reveals novel genetic associations with COVID-19 severity

**DOI:** 10.1101/2021.08.13.21261910

**Authors:** Kenneth E. Westerman, Joanna Lin, Magdalena Sevilla-Gonzalez, Beza Tadess, Casey Marchek, Alisa K. Manning

## Abstract

Increasing evidence indicates that specific genetic variants influence the severity of outcomes after infection with COVID-19. However, it is not clear whether the effect of these genetic factors is independent of the risk due to more established non-genetic demographic and metabolic risk factors such as male sex, poor cardiometabolic health, and low socioeconomic status. We sought to identify interactions between genetic variants and non-genetic risk factors influencing COVID-19 severity via a genome-wide interaction study in the UK Biobank. Of 378,051 unrelated individuals of European ancestry, 2,402 were classified as having experienced severe COVID-19, defined as hospitalization or death due to COVID-19. Exposures included sex, cardiometabolic risk factors (obesity and type 2 diabetes [T2D], tested jointly), and multiple deprivation index. Multiplicative interaction was tested using a logistic regression model, conducting both an interaction test and a joint test of genetic main and interaction effects. Five independent variants reached genome-wide significance in the joint test, one of which also reached significance in the interaction test. One of these, rs2268616 in the *PGF* gene, showed stronger effects in males and in individuals with T2D. None of the five variants showed effects on a similarly-defined phenotype in a lookup in the COVID-19 Host Genetics Initiative. These results reveal potential additional genetic loci contributing to COVID-19 severity and demonstrate the value of including non-genetic risk factors in an interaction testing approach for genetic discovery.

## Introduction

Epidemiological research has uncovered multiple risk factors for COVID-19 severity, including sex, metabolic conditions such as type 2 diabetes and obesity, and socioeconomic status. Male sex is independently associated with higher mortality and worse COVID-19 outcomes (Palaiodimos et al., 2020; Park et al., 2020). Cardiometabolic conditions, such as Type 2 diabetes (T2D) and obesity, are also associated with increased COVID-19 susceptibility and severity (Barron et al., 2020; Zhu et al., 2020). Additionally, associated comorbidities of obesity, such as deregulated immune response, chronic inflammation, metabolic dysfunction, and compromised cilia on airway epithelial cells may put individuals at higher risk of severe COVID-19(Ritter et al., 2020). Minoritized communities are disproportionately impacted by of COVID-19 and may be predisposed to worse conditions due to environmental factors, limited healthcare access, and other societal factors (Tai et al., 2021). Furthermore, housing and neighborhood density and increased work-related exposure may put low-income groups at higher risk (Burström & Tao, 2020). Additionally, the greater prevalence of underlying chronic conditions among individuals with lower socioeconomic status puts this group at greater risk of severe outcomes.

Genetic investigations, such as that from the Host Genetics Initiative (HGI) consortium, have demonstrated that specific genomic regions are associated with COVID-19 severity. The HGI global meta-analysis identified 13 genome-wide significant loci, 9 of which were associated with increased risk of severe symptoms for hospitalized COVID (Ganna, 2021). Several loci were further associated with interstitial lung disease and autoimmune and inflammatory diseases, possibly predisposing individuals to greater immune response and worse outcomes.

It is not clear whether genetic factors impact the relationship between these key risk factors and COVID-19 severity, or whether these interactions can uncover novel genetic loci impacting this outcome. We sought to understand the interactions between genetic variants and previously reported risk factors, in order to gain novel understanding of the underlying mechanisms impacting COVID-19 severity and add an important dimension to the current epidemiological literature on COVID-19. We undertook a series of three genome-wide gene-environment interaction studies in the UK Biobank, while conducting both interaction effect tests and joint tests of genetic main and interaction effects. The “environmental” exposures included sex, cardiometabolic health (obesity and type 2 diabetes status), and social determinants of health (as quantified by the multiple deprivation index). The binary outcome was severe COVID-19 (as defined by hospitalization or death due to COVID-19) while the rest of the population was used as a control group. Using GxE analyses and GWAS post-processing methods, we found 5 genome-wide significant loci that provide insight into the biological mechanisms of severe COVID-19 outcomes.

## Results

The UKB population is described in Table 1, with subjects categorized into those having experienced severe COVID-19 (hospitalization or death from COVID-19; see Methods) and the remaining population (regardless of infection status). While the overall population had a greater proportion of females, cases were more likely to be male (54% vs. 46% in controls, p = 2.3×10^−11^). Cases also had a greater prevalence of T2D (p = 8.2×10^−48^), higher BMI (p = 6.7×10^−62^) and higher MDI (p = 9.7×10^−45^).

**Table 1.** Population characteristics stratified by COVID severity (Total N=378,051) Characteristics of European ancestry samples from the UK Biobank cohort. We present the mean and standard deviation for continuous covariates, percentage of the sample for dichotomous covariates, and p-value for association with severe COVID-19 (*t*-test or Chi-square test for continuous and binary traits, respectively).

| Table 1. Population characteristics stratified by COVID severity<br>(Total N=378,051) |  |  |  |  |
| --- | --- | --- | --- | --- |
|  | Overall | Control<br>(N=375,649) | Severe COVID<br>(N=2,402) | P value |
| **Age**<br>**(years)** | 56.73 (8.02) | 56.7 (8) | 57.9 (8.6) | $2.3 \times 10^{-11}$ |
| **Sex**<br>**(Male)** | 46% | 46% | 54% | $9.4 \times 10^{-17}$ |
| **Body**<br>**Mass**<br>**Index**<br>**(kg/m<sup>2</sup>)** | 27.37 (4.76) | 27.4 (4.8) | 29.3 (5.4) | $6.7 \times 10^{-62}$ |
| **Type 2**<br>**Diabetes** | 4% | 4% | 10% | $8.2 \times 10^{-48}$ |
| **Multiple**<br>**Deprivat**<br>**ion**<br>**Index** | 16.9 (13.5) | 16.8 (13.5) | 22.1 (16.5) | $9.7 \times 10^{-45}$ |

We conducted a GWIS for each of the following exposures: sex, cardiometabolic traits (BMI and T2D, tested jointly), and MDI. Top index variants after pruning are displayed in Suppl. Tables S2-4. Across all scans, five variants (rs2268616, rs182113773, rs148793499, rs11115199, and chr2:218260234) passed a genome-wide significance (GWS) threshold (p < 5×10^−8^) in the joint test. One of these five (rs11115199) was additionally found to be GWS in the cardiometabolic (CM) interaction test (Figure 2; Table 2). Two of these variants (rs148793499, rs11115199) passed a study-wide significance threshold (p < 5×10^−8^/ 3 exposures = 1.6×10^−8^). No variants passed the GWS threshold in the MDI analysis. Of the five variants, a GWS marginal effect was identified for only rs2268616 (p=1.08×10^−8^) and rs182113773 (p=1.39×10^−8^). This result shows that the joint test discovered variants that would not have been found via a standard GWAS in this population.

**Table 2:** Genome-wide significant associations from interaction and joint tests. **A**. Sex interaction and joint tests. **B**. Cardiometabolic interaction and joint tests.

| RSID | location | Effect Allele | Non-Effect Allele | Eff_Allele_Freq | Interaction p-value | Joint p-value | OR interaction | OR combined | OR in males | OR in females |
| --- | --- | --- | --- | --- | --- | --- | --- | --- | --- | --- |
| rs2268616 | 14:75419444 | G | A | 0.018 | 0.14 | $2.7 \times 10^{-8}$ | 1.2 [0.87-1.7] | 1.6 [1.4-1.9] | 1.8 [1.4-2.2] | 1.4 [1.1-1.9] |
| 2:218260234_AC_A | 2:218260234 | A | AC | 0.026 | 0.00013 | $3.0 \times 10^{-8}$ | 1.7 [1.2-2.4] | 1.4 [1.2-1.7] | 1.8 [1.5-2.2] | 1.0 [0.78-1.3] |

B.
| RSID | location | Effect Allele | Non-Effect Allele | Eff_Allele_Freq | Interaction p-value | Joint p-value | OR combined | OR in no_t2d | OR in T2D | OR in no_obesity | OR in obesity |
| --- | --- | --- | --- | --- | --- | --- | --- | --- | --- | --- | --- |
| rs148793499 | 18:58314588 | C | T | 0.010 | $8.4 \times 10^{-6}$ | $1.3 \times 10^{-8}$ | 1.6 [1.3-2.03] | 1.6 [1.2-2.0] | 2.0 [1.0-3.9] | 1.2 [0.83-1.7] | 2.4 [1.7-3.3] |
| rs11115199 | 12:82510665 | T | G | 0.020 | $1.4 \times 10^{-8}$ | $4.8 \times 10^{-8}$ | 0.91 [0.74-1.1] | 0.72 [0.56-0.92] | 2.6 [1.7-3.9] | 1.0 [0.74-1.3] | 0.85 [0.59-1.2] |
| rs182113773 | 7:20239837 | A | C | 0.015 | 0.053 | $2.7 \times 10^{-8}$ | 1.7 [1.4-2.1] | 1.6 [1.3-2.0] | 2.6 [1.6-4.3] | 1.6 [1.3-2.1] | 1.9 [1.4-2.6] |
| rs2268616 | 14:75419444 | G | A | 0.018 | 0.26 | $3.9 \times 10^{-8}$ | 1.6 [1.4-1.9] | 1.6 [1.3-1.9] | 2.0 [1.2-3.3] | 1.8 [1.5-2.2] | 1.4 [1.0-1.9] |

**Figure 1:**
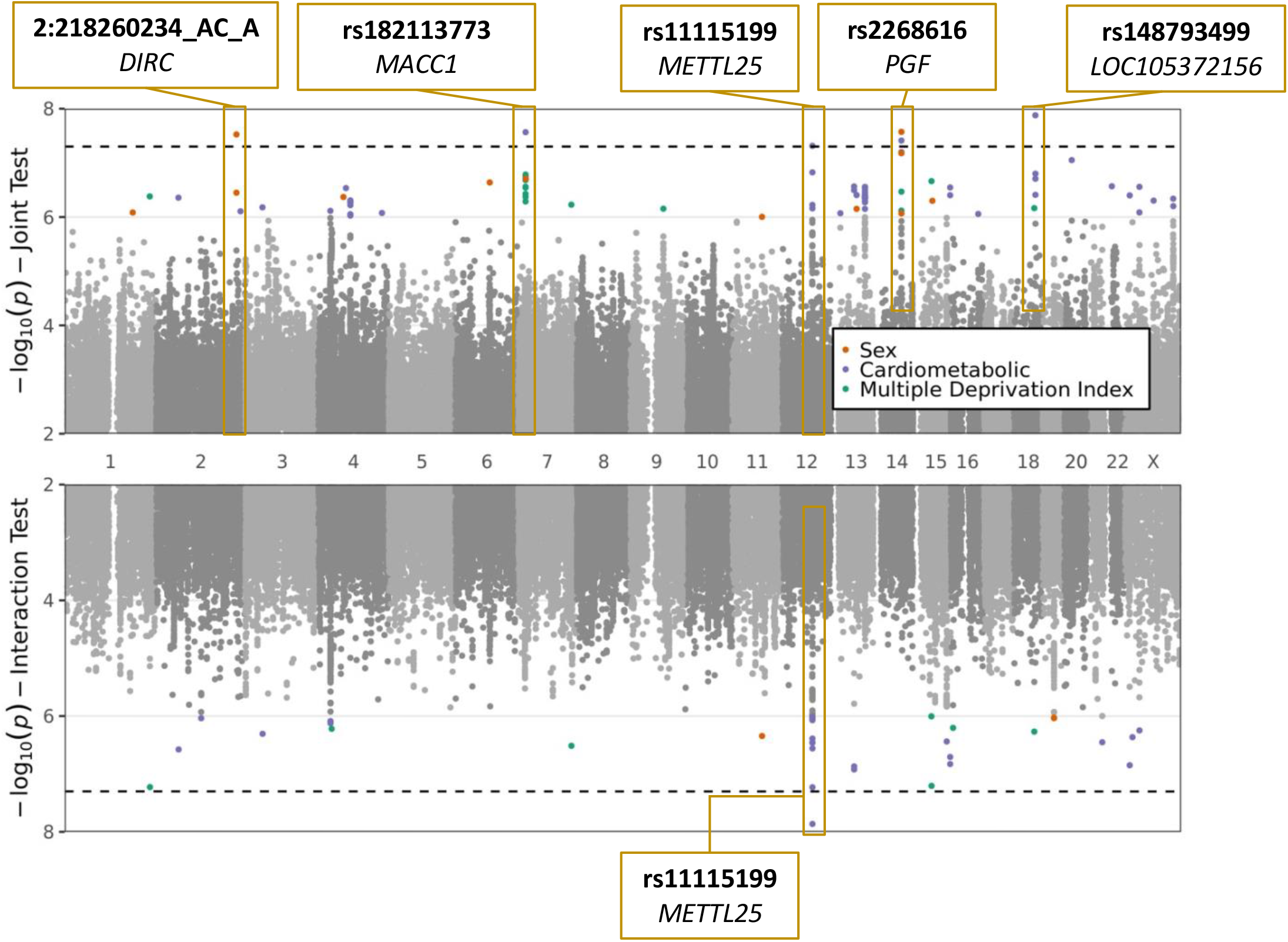
Plots of sex, cardiometabolic, and MDI joint and interaction tests. The upper plot shows negative logarithm of joint p-values in a test of main and interaction effects, while the lower plot shows negative logarithm of the interaction test p-values. X-axis corresponds to genomic position. Genome-wide significant loci are labeled with the most significant variant at the locus and the annotated to genes based on proximity (*DIRC, MACC1, PGF, LOC105372156*) or eQTL relationships (*METTL25*).

**Figure 2:**
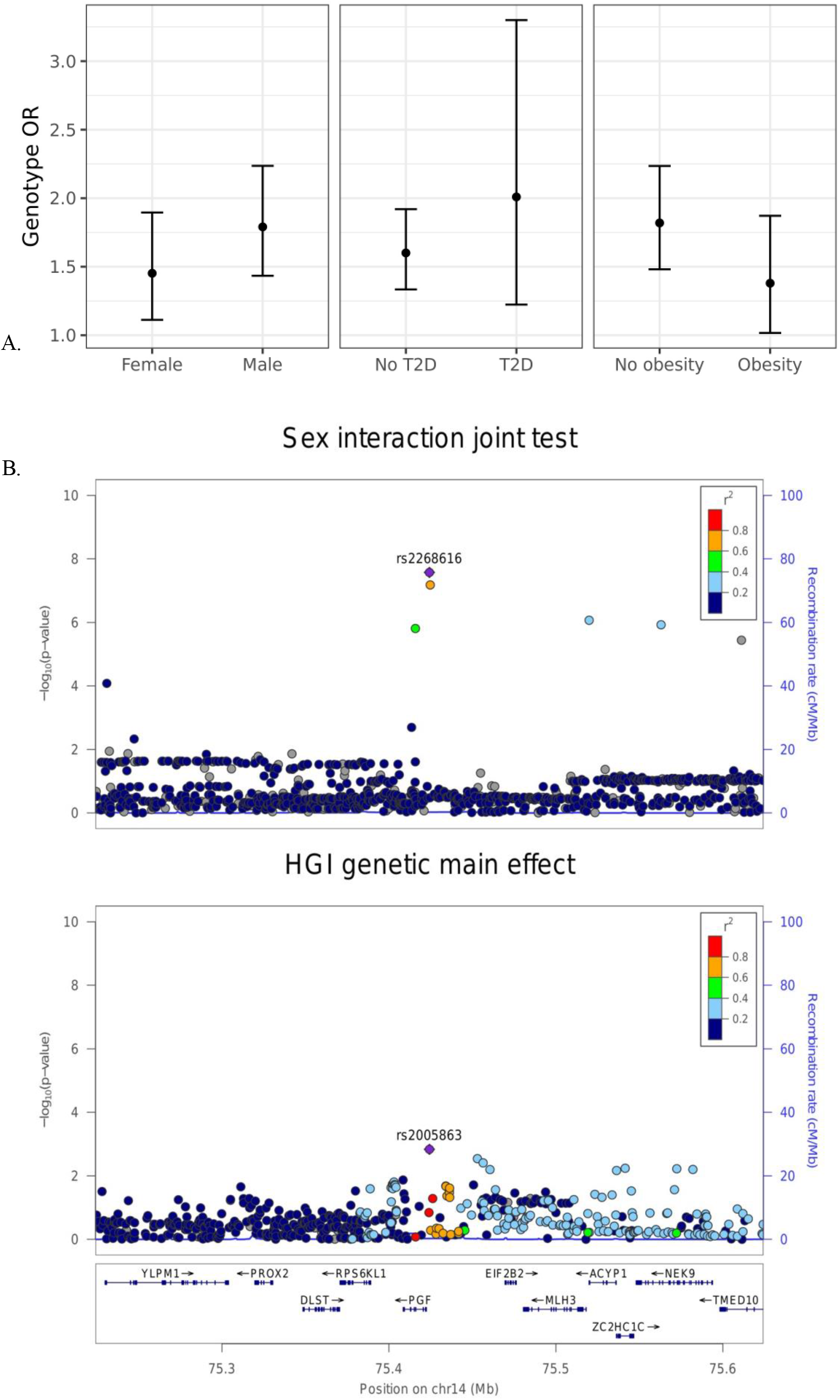
Inspection of the sex-rs2268616 interaction effect. (A) Stratified genetic effects on severe COVID-19 after adjustment for the primary set of covariates. Y-axis indicates the estimated odds ratio for severe COVID-19 per alternate allele. Strata are defined by (left to right): sex (male or female), T2D status, and obesity (BMI less than or greater than 30). (B) Regional association plots showing association signals from this analysis (sex joint test, top panel) and HGI B2 phenotype meta-analysis (genetic main effects with UKB omitted, bottom panel).

These five GWS variants were compared to genetic main effects from the HGI meta-analysis (with UKB omitted) testing the equivalent “B2” phenotype (hospitalized COVID-19 vs. population). A significant genetic main effect would constitute a partial replication of the joint test (genetic plus interaction effect) hypothesis. Neither of the two variants directly tested in the HGI meta-analysis showed nominal replication (both p > 0.05). For the remaining three, neither the variants nor close genetic proxies (r^2^ > 0.5 using European-based linkage disequilibrium patterns) were available in the HGI dataset.

Next, we explored these top variants and interactions to understand their potential biological function. One variant of interest, rs2268616 (MAF=0.018), was genome-wide significant in the joint sex analyses (p=2.67×10^−8^) and joint cardiometabolic diseases (p=3.87×10^−8^). This variant sits in an intron of the placental growth factor (PGF) gene and is associated with testosterone in GWAS analyses. It is also a putative enhancer in lung and other tissues, and is an eQTL for EIF2B2 (a gene in a family of proteins that regulate viral mRNA translation) in whole blood. However, colocalization analysis using whole blood eQTL statistics from eQTLGen Consortium did not support the hypothesis of a shared causal variant with either PGF or EIF2B (posterior probabilities <0.1%). Sex-stratified analysis showed a stronger genetic effect in males (OR [95% CI] =1.79 [1.43-2.24]) compared to females (OR=1.45 [1.11-1.9]), as shown in Figure 3A. T2D-stratified tests also showed a greater genetic effect on severe COVID-19 in individuals with T2D (OR=2.01 [1.22-3.32]) compared to those without T2D (OR=1.6 [1.33-1.9).

The additional GWS variants also indicated genetic effects on COVID-19 severity mediated through interaction effects. rs182113773 was found in the cardiometabolic joint test (p = 2.71×10^−8^) and found in the intron for MACC1. This variant sits in an enhancer within neutrophils, monocytes, and B cells and has a RegulomeDB score of 0.59, suggesting a regulatory role in transcription. Variant chr2:218260234 was found in the sex analysis joint test (p = 2.99×10^−8^, MAF=0.025). Stratified analysis for this variant demonstrated a strong genetic effect in males (OR=1.8 [1.47-2.19]) that was not found in females (OR = 1.03 [0.779-1.35]). In addition, rs11115199 is an intergenic variant that was identified in both the cardiometabolic interaction and joint tests (respectively, p = 1.37×10^−8^ & 4.85×10^−8^, MAF = 0.02). rs11115199 is an eQTL for *METTL25* based on the GTEx database and has modest associations with cardiometabolic traits (positive with weight and BMI-adjusted T2D, negative with obesity). Finally, rs148793499 was identified in the cardiometabolic joint test (p=1.8×10^−10^, MAF=0.01). Stratified genetic effects showed more pronounced associations in obesity (OR = 2.36 [1.7-3.27]) with a similar but weaker pattern for T2D (OR = 2.01 [1.03-3.93]).

## Discussion

Exploring the interplay of genetics and sex offers novel understanding of the underlying mechanisms impacting COVID-19 severity and adds an important dimension to the current epidemiological literature on COVID-19. In this genome-wide gene-environment interaction analysis, we found five significant genomic regions (p<5×10^−8^) that interact with well-established risk factors to influence COVID-19 severity.

Sex-dimorphic transcripts and hormones, as well as differences in environmental factors between the sexes, contribute to differential immune responses between sexes (Klein & Flanagan, 2016) and may mediate the established association of male sex with greater COVID-19 severity. In our analysis, rs2268616 was statistically significant in the joint analyses for sex and cardiometabolic diseases (p<5×10^−8^). This variant has been associated with testosterone and placental growth factor gene in GWAS analyses, suggesting that this variant interacts with sex to mediate worse COVID-19 outcomes. Interestingly, this variant is also an eQTL for *EIF2B2*, a gene within a family of proteins that mediate viral mRNA translation. Moreover, prior studies have found an increased risk of death and significantly increased levels of inflammatory markers in male COVID-19 positive hospitalized patients compared with women (Lau et al., 2021). The *EIF2B2* variant is linked to a strong transcription chromatin state in the cells of the lung, spleen, and B-cells, perhaps mediating the robust inflammatory response in males that is associated with worse COVID outcomes. Furthermore, rs2268616 sits within an enhancer in lung tissue, suggesting a role of this variant on transcription and respiratory complications after SARS-CoV-2 infection. Rs2268616 also shows a modest positive association with coronary artery disease and negative association with HOMA-B based on lookups in the Type 2 Diabetes Knowledge Portal (https://t2d.hugeamp.org/), indicating a potential influence on metabolic traits in general. Our findings suggest that this genetic variant may modify the relationship between biological differences and associated worse COVID-19 outcomes primarily through regulating viral RNA clearance immune response and lung cell transcription.

Comorbidities associated with cardiometabolic health such as obesity and T2D have been implicated in mediating worse COVID-19 outcomes (Ritter et al., 2020). Our findings show four variants that were genome-wide significant in our cardiometabolic joint tests: rs182113773, rs11115199, rs148793499 and rs2268616. Located within the intron for *MACC1*, a gene associated with BMI-adjusted waist circumference and BMI-adjusted waist-hip ratio, rs182113773 is an enhancer within neutrophils, monocytes, and B cells. This variant also has high gene expression in EBV-transformed lymphocytes and is a likely regulatory variant (RegulomeDB score of 0.59), which further suggests that the interaction of this variant with cardiometabolic health has a regulatory role on immune response. Studies found that increased neutrophil count in T2D groups are associated with clinical severity and may mediate the positive association between T2D and COVID-19 severity (Zhu et al., 2020). Thus, this *MACC1* variant may be interacting with cardiometabolic health to mediate greater COVID-19 severity. Furthermore, obese adipose tissues overexpress receptors and proteases that enable the entry of SARS-CoV-2, possibly contributing to the severe inflammation and immune response of individuals with obesity (Ritter et al., 2020).

Alongside decreased immune response mediated by testosterone, rs2268616 may also play a role in the deflated immune response seen in cases with cardiometabolic disease status. This variant has a positive association with coronary artery disease and a negative association with HOMA-B (a method that assesses β-cell function from basal fasting glucose and insulin). For cardiometabolic diseases, well controlled blood glucose and smaller glycemic variability have been associated with lower mortality during hospitalization due to COVID-19 (Zhu et al., 2020). Therefore, this variant may help explain the COVID-19 biology that increases the risk for individuals with T2D. Interactions between these genetic factors and deregulated immune response, chronic inflammation, metabolic dysfunction, and other comorbidities of obesity and T2D may be placing individuals at greater risk for worse outcomes of COVID-19.

Beyond rs2268616, other genome-wide significant variants identified in the cardiometabolic analysis are of potential biological interest. The rs11115199 variant is an eQTL for *METTL25*, a gene that has known genetic links to BMI but which has minimal transcription in memory T cells and B cells. Thus, this locus may instead modify immune response via interactions with obesity. Meanwhile, the genetic effect of rs148793499 appears to be most directly modulated by metabolic status; in stratified cardiometabolic tests, the variant showed strong effects in individuals with T2D but no major differences in effect in individuals with obesity.

Our analysis focusing on social determinants of health did not identify any significant variants. This may be a function of the noise associated with the MDI measurement and the difficulty in using this measurement to represent social determinants of health in a large diverse population. One study leveraged an Index of Multiple Deprivation and Income Deprivation Affecting Older People Index to show higher incidence of COVID-19 related deaths in the most deprived quartiles (Bach-Mortensen & Degli Esposti, 2021). We subsetted our sample to participants from England to reduce heterogeneity, but this reduced the sample size (by 16.5%; 2,007 vs. 2,402 cases) and thus statistical power available for the MDI analysis. Additionally, there may simply be little signal to uncover: the effects of genetics and social determinants of health on COVID-19 severity may be approximately independent.

The results of this study may be limited due to linkage disequilibrium and heterogeneity caused by geographic location within our sample population. The case definition allows us to identify variants associated with severity, however these results need to be taken with caution given the possibility of collider bias. Analyzing UK Biobank data, the participants tested for COVID-19 were highly selected for a range of genetic, behavioral, cardiovascular, demographic, and anthropometric traits (Griffith et al., 2020). By subsetting our dataset to individuals of European ancestry, we reduce the heterogeneity but face a limited sample size. Nonetheless, the use of interaction analysis allowed us to uncover novel variants: the GEM marginal p-value did not pass the genome-wide significance threshold for three of the five variants, meaning that these variants would not have been detected via a standard GWAS in this population.

Our findings suggest that gene-environment interaction effects contribute to the differences in COVID-19 severity. Sex-associated differences in immune response and cardiometabolic disease comorbidities that deregulate immune response may interact with the identified genetic variants and put individuals at higher risk for worse outcomes of COVID-19. Future studies investigating the stratified effects of sex, T2D and BMI, and social determinants of health on COVID-19 susceptibility, as well as similar analysis with a wider array of ancestries, may further reveal underlying the genetic interaction effects that place individuals at higher risk.

## Methods

### UK Biobank Dataset

The UK Biobank (UKB) is a population-based cohort including over 500,000 individuals living in England, Wales, and Scotland. The sub-population of interest for this study included unrelated individuals of European ancestry in order to minimize genetic heterogeneity. Sample sizes varied depending on available phenotypes across these populations. COVID-19 test results were downloaded from the UKBB data portal on January 1, 2020. The severe COVID-19 phenotype for was defined as laboratory confirmed SARS-CoV-2 infection plus hospitalized COVID-19, with the rest of the population serving as controls versus the rest of the population. This definition was designed to mirror that of the “B2” phenotype used by the COVID-19 Host Genetics Initiative team (Ganna, 2021) (COVID-19 Host Genetics Initiative, 2020) and is outlined in Supp. Fig. 1. Genotype preprocessing was primarily performed centrally by the UKB with filters at the marker and sample level (Bycroft et al., 2018). Genotypes were further subsetted to common variants (minor allele frequency > 0.05) for analysis.

### Exposures of Interest

Risk factors used as exposures were measures of genetically-determined sex, cardiometabolic health, and social determinants of health (SDH). For cardiometabolic measures, BMI was used as a measure of obesity and T2D status was determined based on self-reported medical history and medication use (“probable” or “possible” algorithmic definitions described by Eastwood an colleagues (Eastwood et al., 2016). BMI and T2D were tested jointly, and then individually as a sensitivity analysis. The multiple deprivation index (MDI) was used as a measure of social determinants of health (SDH). The MDI is composed of metrics including economic stability, physical environment, and education; details can be found at https://biobank.ndph.ox.ac.uk/ukb/label.cgi?id=76. For the MDI analyses only, only the subset of the population living in England was used in order to reduce heterogeneity.

### Statistical analysis

A genome-wide scan was performed based on a logistic regression model including gene-environment interaction terms :

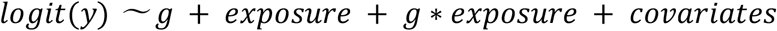

Y was the binary severe COVID-19 indicator (defined above). The three genome-wide scans used the following exposures: sex, cardiometabolic conditions (BMI and T2D), and MDI. For the cardiometabolic conditions, two environmental terms and two interaction terms were tested jointly. To test T2D exposure effect, GxT2D interaction obese and non-obese stratified analyses were run. Covariates included age, five genetic principal components, and sex. Genome-wide analysis was conducted using GEM v1.2 (Westerman et al., 2020) with robust standard errors. For each variant, two statistical tests were derived: an interaction test and a joint test of the interaction term(s) plus the genetic main effect.

Interaction and joint analyses were conducted on the Terra cloud platform. Phenotype definitions and population summaries were created in interactive Jupyter notebooks with an R 3.6 kernel. GWIS analyses were submitted as workflows using a Workflow Description Language (WDL) script implementing GEM. Post-GWIS summarization and visualizations were created in a separate Jupyter notebook. These notebooks can be viewed on GitHub (https://github.com/manning-lab/ukb-covid-gxe).

### Variant Biology Investigation

Top variants were further investigated for trait associations, eQTLs, and linkage disequilibrium using dbSNP (NCBI), PhenoScanner (Kamat et al., 2019; Staley et al., 2016), RegulomeDB (Boyle et al., 2012), Type 2 Diabetes Knowledge portal (https://t2d.hugeamp.org/), and LDlink (Myers et al., 2020). Colocalization between interactions and eQTLs was performed using the *coloc* package (Giambartolomei et al., 2014) along with blood-based eQTL summary statistics from the eQTLGen Consortium (Võsa et al., 2018)

## Data Availability

Publicly available data from the UK Biobank study was analyzed in this study. The datasets are available to researchers through an open application via https://www.ukbiobank.ac.uk/register-apply/.

## Author Contributions

KEW – Conceptualization, Data curation, UK Biobank Analysis, Software, Writing

JL – Phenotype Definition, Summary Statistics Analysis, Writing

MSG – Phenotype Definition, Writing, Manuscript Review and Editing

BT – Phenotype Definition, Manuscript Review and Editing

CM – Manuscript Review and Editing

AKM – Conceptualization, Funding, Supervision, Manuscript Review and Editing

## Acknowledgments

JL was funded by the Massachusetts General Hospital COVID Corps Biomedical Research Internship Program. KEW, MSG, BT, CM and AKM were funded by NIH R01 HL145025. The UK Biobank analysis was performed at the Broad Institute under UK Biobank application #27892. While KEW, AKM, and MSG are employees of Mass General Brigham (MGB), this work was not conducted in their capacity as MGB employees.

## References

Bach-Mortensen, A. M., & Degli Esposti, M. (2021). Is area deprivation associated with greater impacts of COVID-19 in care homes across England? A preliminary analysis of COVID-19 outbreaks and deaths. Journal of Epidemiology and Community Health, jech-2020-215039. https://doi.org/10.1136/jech-2020-215039

Barron, E., Bakhai, C., Kar, P., Weaver, A., Bradley, D., Ismail, H., Knighton, P., Holman, N., Khunti, K., Sattar, N., Wareham, N. J., Young, B., & Valabhji, J. (2020). Associations of type 1 and type 2 diabetes with COVID-19-related mortality in England: a whole - population study. The Lancet Diabetes & Endocrinology, 8(10), 813–822. https://doi.org/10.1016/S2213-8587(20)30272-2

Boyle, A. P., Hong, E. L., Hariharan, M., Cheng, Y., Schaub, M. A., Kasowski, M., Karczewski, K. J., Park, J., Hitz, B. C., Weng, S., Cherry, J. M., & Snyder, M. (2012). Annotation of functional variation in personal genomes using RegulomeDB. Genome research, 22(9), 1790–1797. https://doi.org/10.1101/gr.137323.112

Burström, B., & Tao, W. (2020). Social determinants of health and inequalities in COVID-19. European Journal of Public Health, 30(4), 617–618. https://doi.org/10.1093/eurpub/ckaa095

Bycroft, C., Freeman, C., Petkova, D., Band, G., Elliott, L. T., Sharp, K., Motyer, A., Vukcevic, D., Delaneau, O., O’Connell, J., Cortes, A., Welsh, S., Young, A., Effingham, M., McVean, G., Leslie, S., Allen, N., Donnelly, P., & Marchini, J. (2018). The UK Biobank resource with deep phenotyping and genomic data. Nature, 562(7726), 203–209. https://doi.org/10.1038/s41586-018-0579-z

Eastwood, S. V., Mathur, R., Atkinson, M., Brophy, S., Sudlow, C., Flaig, R., de Lusignan, S., Allen, N., & Chaturvedi, N. (2016). Algorithms for the Capture and Adjudication of Prevalent and Incident Diabetes in UK Biobank. PLOS ONE, 11(9), e0162388. https://doi.org/10.1371/journal.pone.0162388

Ganna, A. (2021). Mapping the human genetic architecture of COVID-19 by worldwide meta-analysis. medRxiv, 2021.2003.2010.21252820. https://doi.org/10.1101/2021.03.10.21252820

Giambartolomei, C., Vukcevic, D., Schadt, E. E., Franke, L., Hingorani, A. D., Wallace, C., & Plagnol, V. (2014). Bayesian Test for Colocalisation between Pairs of Genetic Association Studies Using Summary Statistics. PLOS Genetics, 10(5), e1004383. https://doi.org/10.1371/journal.pgen.1004383

Griffith, G. J., Morris, T. T., Tudball, M., Herbert, A., Mancano, G., Pike, L., Sharp, G. C., Palmer, T. M., Smith, G. D., Tilling, K., Zuccolo, L., Davies, N. M., & Hemani, G. (2020). Collider bias undermines our understanding of COVID-19 disease risk and severity. medRxiv, 2020.2005.2004.20090506. https://doi.org/10.1101/2020.05.04.20090506

Kamat, M. A., Blackshaw, J. A., Young, R., Surendran, P., Burgess, S., Danesh, J., Butterworth, A. S., & Staley, J. R. (2019). PhenoScanner V2: an expanded tool for searching human genotype–phenotype associations. Bioinformatics, 35(22), 4851–4853. https://doi.org/10.1093/bioinformatics/btz469

Klein, S. L., & Flanagan, K. L. (2016). Sex differences in immune responses. Nature Reviews Immunology, 16(10), 626–638. https://doi.org/10.1038/nri.2016.90

Lau, E. S., McNeill, J. N., Paniagua, S. M., Liu, E. E., Wang, J. K., Bassett, I. V., Selvaggi, C. A., Lubitz, S. A., Foulkes, A. S., & Ho, J. E. (2021). Sex differences in inflammatory markers in patients hospitalized with COVID-19 infection: Insights from the MGH COVID-19 patient registry. PLOS ONE, 16(4), e0250774. https://doi.org/10.1371/journal.pone.0250774

Myers, T. A., Chanock, S. J., & Machiela, M. J. (2020). LDlinkR: An R Package for Rapidly Calculating Linkage Disequilibrium Statistics in Diverse Populations [Technology and Code]. Frontiers in Genetics, 11(157). https://doi.org/10.3389/fgene.2020.00157

Palaiodimos, L., Kokkinidis, D. G., Li, W., Karamanis, D., Ognibene, J., Arora, S., Southern, W. N., & Mantzoros, C. S. (2020). Severe obesity, increasing age and male sex are independently associated with worse in-hospital outcomes, and higher in-hospital mortality, in a cohort of patients with COVID-19 in the Bronx, New York. Metabolism, 108, 154262. https://doi.org/10.1016/j.metabol.2020.154262

Park, R., Chidharla, A., Mehta, K., Sun, W., Wulff-Burchfield, E., & Kasi, A. (2020). Sex-bias in COVID-19-associated illness severity and mortality in cancer patients: A systematic review and meta-analysis. EClinicalMedicine, 26, 100519. https://doi.org/10.1016/j.eclinm.2020.100519

Ritter, A., Kreis, N.-N., Louwen, F., & Yuan, J. (2020). Obesity and COVID-19: Molecular Mechanisms Linking Both Pandemics. International Journal of Molecular Sciences, 21(16), 5793. https://www.mdpi.com/1422-0067/21/16/5793

Staley, J. R., Blackshaw, J., Kamat, M. A., Ellis, S., Surendran, P., Sun, B. B., Paul, D. S., Freitag, D., Burgess, S., Danesh, J., Young, R., & Butterworth, A. S. (2016). PhenoScanner: a database of human genotype–phenotype associations. Bioinformatics, 32(20), 3207–3209. https://doi.org/10.1093/bioinformatics/btw373

Tai, D. B. G., Shah, A., Doubeni, C. A., Sia, I. G., & Wieland, M. L. (2021). The Disproportionate Impact of COVID-19 on Racial and Ethnic Minorities in the United States. Clin Infect Dis, 72(4), 703–706. https://doi.org/10.1093/cid/ciaa815

Võsa, U., Claringbould, A., Westra, H.-J., Bonder, M. J., Deelen, P., Zeng, B., Kirsten, H., Saha, A., Kreuzhuber, R., Kasela, S., Pervjakova, N., Alvaes, I., Fave, M.-J., Agbessi, M., Christiansen, M., Jansen, R., Seppälä, I., Tong, L., Teumer, A., Schramm, K., Hemani, G., Verlouw, J., Yaghootkar, H., Sönmez, R., Brown, A., Kukushkina, V., Kalnapenkis, A., Rüeger, S., Porcu, E., Kronberg-Guzman, J., Kettunen, J., Powell, J., Lee, B., Zhang, F., Arindrarto, W., Beutner, F., Brugge, H., Dmitreva, J., Elansary, M., Fairfax, B. P., Georges, M., Heijmans, B. T., Kähönen, M., Kim, Y., Knight, J. C., Kovacs, P., Krohn, K., Li, S., Loeffler, M., Marigorta, U. M., Mei, H., Momozawa, Y., Müller-Nurasyid, M., Nauck, M., Nivard, M., Penninx, B., Pritchard, J., Raitakari, O., Rotzchke, O., Slagboom, E. P., Stehouwer, C. D. A., Stumvoll, M., Sullivan, P., Hoen, P. A. C. t., Thiery, J., Tönjes, A., van Dongen, J., van Iterson, M., Veldink, J., Völker, U., Wijmenga, C., Swertz, M., Andiappan, A., Montgomery, G.W., Ripatti, S., Perola, M., Kutalik, Z., Dermitzakis, E., Bergmann, S., Frayling, T., van Meurs, J., Prokisch, H., Ahsan, H., Pierce, B., Lehtimäki, T., Boomsma, D., Psaty, B. M., Gharib, S. A., Awadalla, P., Milani, L., Ouwehand, W., Downes, K., Stegle, O., Battle, A., Yang, J., Visscher, P. M., Scholz, M., Gibson, G., Esko, T., & Franke, L. (2018). Unraveling the polygenic architecture of complex traits using blood eQTL metaanalysis. bioRxiv, 447367. https://doi.org/10.1101/447367

Westerman, K. E., Pham, D. T., Hong, L., Chen, Y., Sevilla-González, M., Sung, Y. J., Sun, Y. V., Morrison, A. C., Chen, H., & Manning, A. K. (2020). GEM: Scalable and flexible gene-environment interaction analysis in millions of samples. bioRxiv, 2020.2005.2013.090803. https://doi.org/10.1101/2020.05.13.090803

Zhu, L., She, Z. G., Cheng, X., Qin, J. J., Zhang, X. J., Cai, J., Lei, F., Wang, H., Xie, J., Wang, W., Li, H., Zhang, P., Song, X., Chen, X., Xiang, M., Zhang, C., Bai, L., Xiang, D., Chen, M. M., Liu, Y., Yan, Y., Liu, M., Mao, W., Zou, J., Liu, L., Chen, G., Luo, P., Xiao, B., Zhang, C., Zhang, Z., Lu, Z., Wang, J., Lu, H., Xia, X., Wang, D., Liao, X., Peng, G., Ye, P., Yang, J., Yuan, Y., Huang, X., Guo, J., Zhang, B. H., & Li, H. (2020). Association of Blood Glucose Control and Outcomes in Patients with COVID-19 and Pre-existing Type 2 Diabetes. Cell Metab, 31(6), 1068-1077.e1063. https://doi.org/10.1016/j.cmet.2020.04.021

