## Supplementary material for "Gene-environment interaction analysis incorporating sex, cardiometabolic diseases, and multiple deprivation index reveals novel genetic associations with COVID-19 severity": Supp. Fig.

**Supplemental Figures**

**Supp. Fig. S1**: Severe COVID-19 phenotype definition flow chart. This definition is based on the “B2” phenotype used by the COVID-19 HGI group.

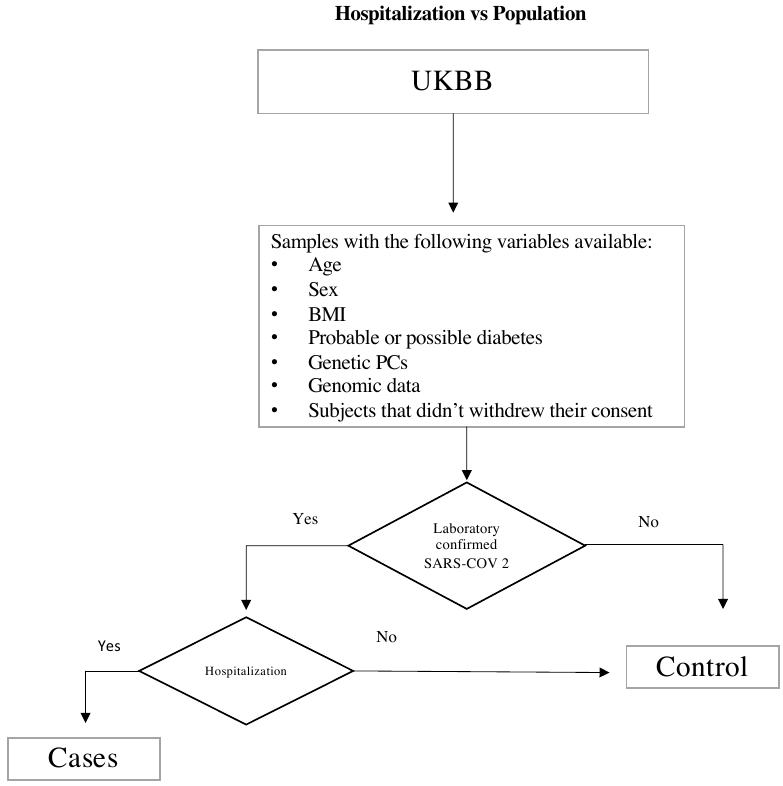

**Supp. Fig. S2**: Sex analysis joint and interaction analysis results. The Manhattan plot displays association test strengths for the joint (top panel) and interaction (bottom panel) tests as a function of genomic position (*x*-axis).

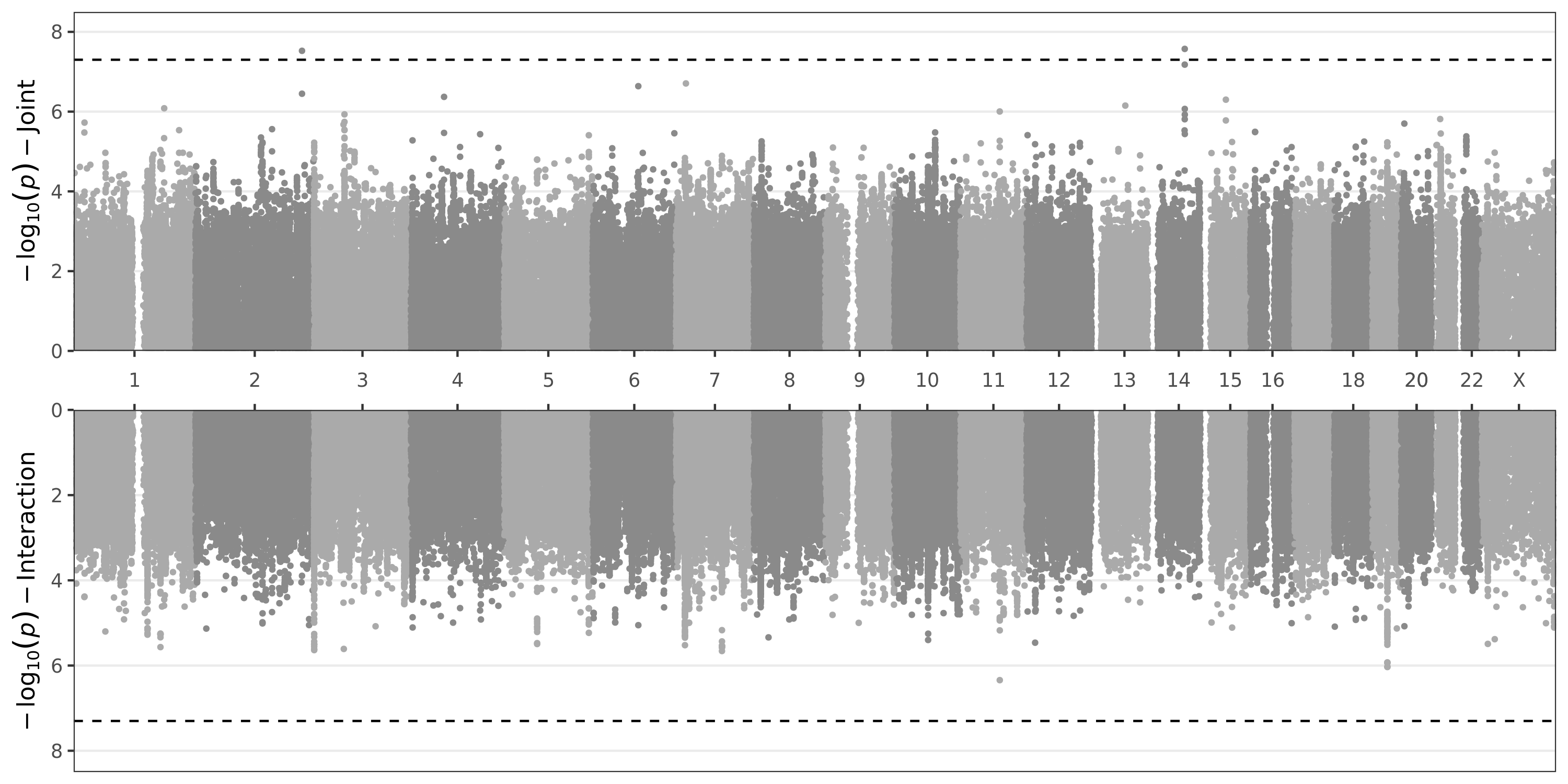

**Supp. Fig. S3**: Cardiometabolic analysis joint and interaction analysis results. The Manhattan plot displays association test strengths for the joint (top panel) and interaction (bottom panel) tests as a function of genomic position (*x*-axis).

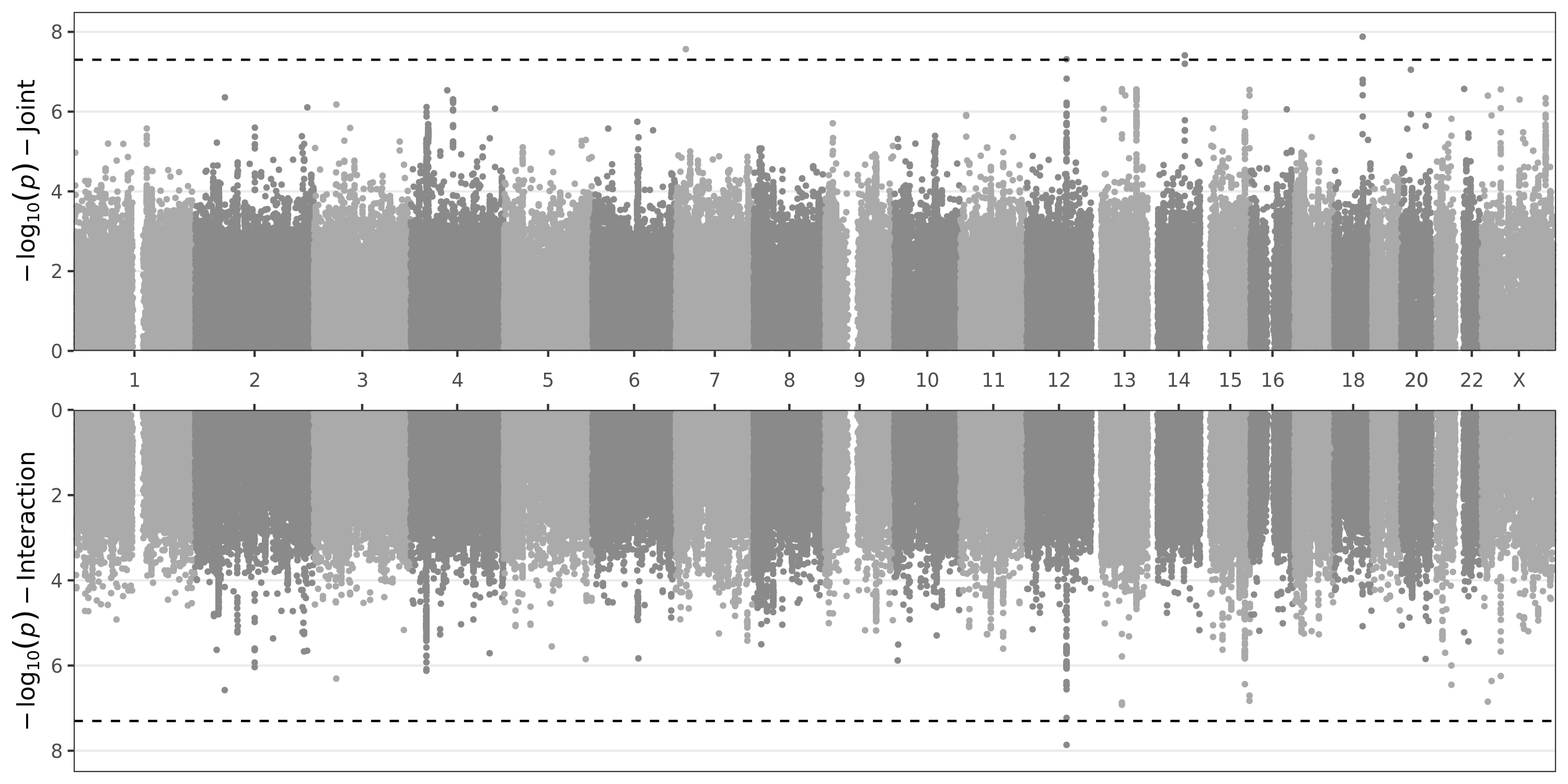

**Supp. Fig. S4**: Multiple deprivation index analysis joint and interaction analysis results. The Manhattan plot displays association test strengths for the joint (top panel) and interaction (bottom panel) tests as a function of genomic position (*x*-axis).

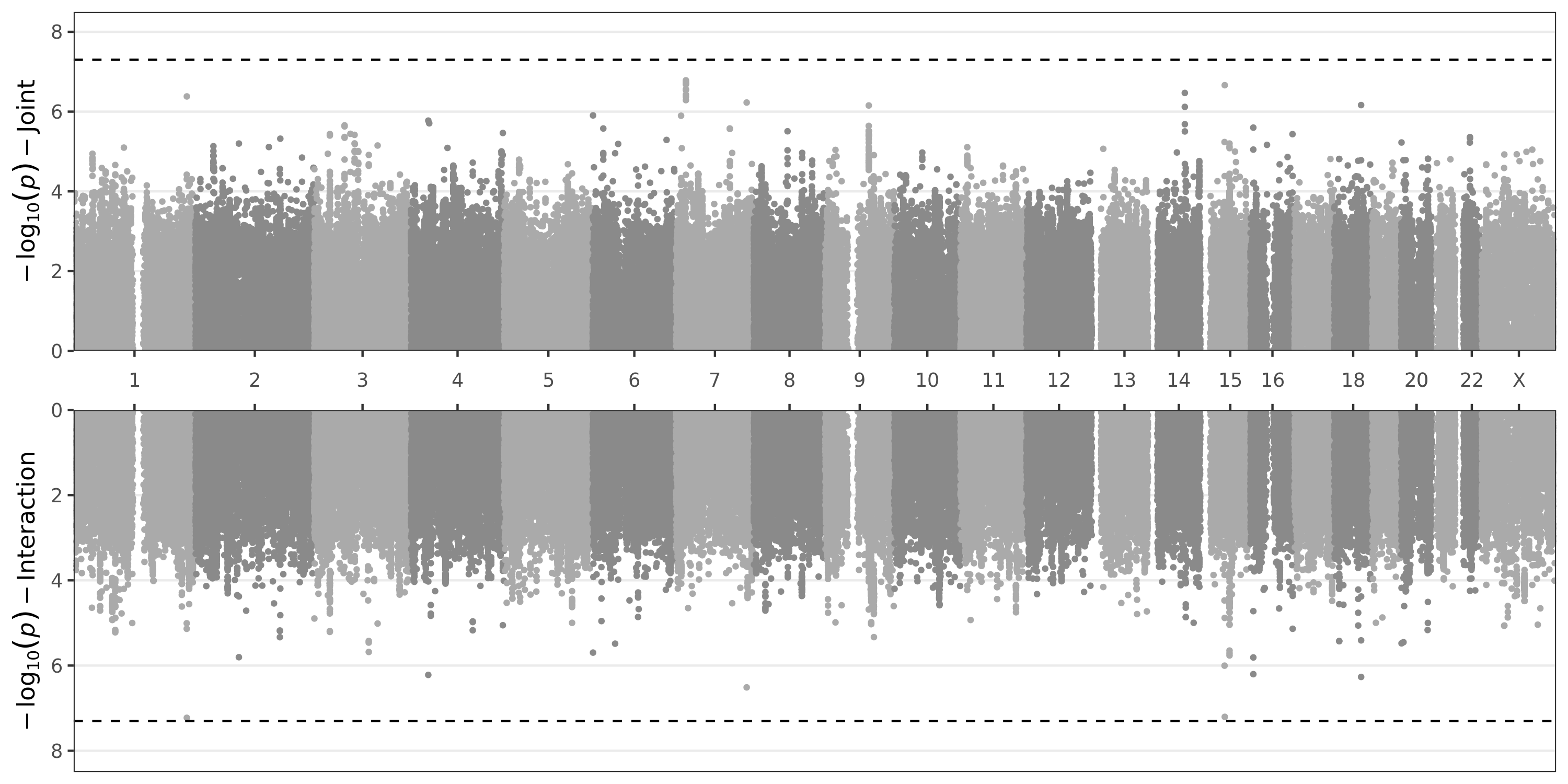
