## Supplementary material for "Gene-environment interaction analysis incorporating sex, cardiometabolic diseases, and multiple deprivation index reveals novel genetic associations with COVID-19 severity": Suppl. Tables

**Supplemental Tables**

**Supp. Table 1:** Population characteristics of European ancestry samples extracted from UKB, stratified by sex.

| **Supp. Table 1. Population characteristics stratified by sex** (n=378,051) | | |
| --- | --- | --- |
| **Sex** | Female  (n= 203,961) | Male  (n= 174,090) |
| **Age** | 56.5 (7.9) | 57 (8.1) |
| **BMI** | 27 (5.1) | 27.8 (4.2) |
| **T2D** | 3% | 6% |
| **MDI** | 16.6 (13.2) | 17.2 (13.8) |

**Supp. Table 2:** Top variants from the sex analysis (joint and interaction tests) along with stratified genetic odds ratios.

| **RSID** | **location** | **Effect Allele** | **Non-Effect Allele** | **Effect Allele**  **Freq** | **GEM Interaction p-value** | **GEM Joint p-value** | **OR interaction** | **OR combined** | **OR (male)** | **OR (female)** |
| --- | --- | --- | --- | --- | --- | --- | --- | --- | --- | --- |
| rs2268616 | 14:75419444 | G | A | 0.018 | 0.139689 | 2.67×10^-8^ | 1.23 [0.868-1.75] | 1.64 [1.38-1.94] | 1.79 [1.43-2.24] | 1.45 [1.11-1.9] |
| 2:218260234_AC_A | 2:218260234 | A | AC | 0.026 | 0.000128 | 2.99×10^-8^ | 1.74 [1.24-2.45] | 1.44 [1.22-1.69] | 1.8 [1.47-2.19] | 1.03 [0.779-1.35] |
| rs182113773 | 7:20239837 | A | C | 0.015 | 0.866041 | 1.96×10^-7^ | 0.98 [0.657-1.45] | 1.73 [1.42-2.1] | 1.71 [1.3-2.23] | 1.75 [1.31-2.34] |
| rs117993077 | 6:93824151 | T | G | 0.012 | 8.86×10^-6^ | 2.3×10^-7^ | 2.67 [1.55-4.57] | 1.47 [1.16-1.85] | 2.06 [1.57-2.7] | 0.78 [0.487-1.24] |
| rs55926550 | 4:68007466 | C | G | 0.012 | 0.002296 | 4.27×10^-7^ | 0.59 [0.388-0.911] | 1.6 [1.3-1.98] | 1.22 [0.882-1.69] | 2.05 [1.55-2.7] |
| rs11232355 | 11:80540380 | G | A | 0.069 | 4.54×10^-7^ | 9.92×10^-7^ | 0.58 [0.463-0.725] | 1.09 [0.972-1.21] | 0.82 [0.694-0.972] | 1.42 [1.22-1.64] |
| rs117234438 | 15:52349607 | G | A | 0.011 | 0.000927 | 5.01×10^-7^ | 1.93 [1.17-3.19] | 1.64 [1.3-2.06] | 2.09 [1.59-2.76] | 1.08 [0.71-1.64] |
| rs796925608 | 13:68788267 | ACTTTGAAAATAAC | A | 0.898 | 0.841366 | 7.06×10^-7^ | 1.02 [0.827-1.26] | 0.76 [0.681-0.839] | 0.76 [0.662-0.879] | 0.75 [0.64-0.872] |
| rs112561874 | 1:185427460 | A | G | 0.016 | 3.54×10^-5^ | 8.24×10^-7^ | 0.47 [0.305-0.723] | 1.42 [1.15-1.74] | 0.94 [0.668-1.32] | 2 [1.53-2.6] |
| rs12461506 | 19:30878810 | C | G | 0.643 | 9.24304×10^-7^ | 5.89398×10^-6^ | 1.35 [1.19-1.52] | 1 [0.943-1.06] | 1.15 [1.06-1.25] | 0.85 [0.783-0.932] |
| rs17066139 | 3:62113696 | T | C | 0.115 | 0.013713 | 1.17×10^-6^ | 1.22 [1.03-1.44] | 1.22 [1.12-1.32] | 1.33 [1.19-1.48] | 1.09 [0.955-1.24] |
| rs114103616 | 21:16998740 | T | A | 0.028 | 0.014151 | 1.53×10^-6^ | 0.7 [0.501-0.983] | 1.47 [1.24-1.74] | 1.23 [0.964-1.58] | 1.77 [1.4-2.22] |
| rs182465934 | 1:22335633 | A | C | 0.028 | 4.16×10^-5^ | 1.88×10^-6^ | 1.93 [1.35-2.77] | 1.3 [1.1-1.54] | 1.67 [1.36-2.05] | 0.87 [0.645-1.17] |
| rs758053125 | 20:6586718 | T | C | 0.050 | 8.37×10^-6^ | 2×10^-6^ | 0.52 [0.379-0.712] | 1.22 [1.04-1.43] | 0.87 [0.688-1.11] | 1.68 [1.36-2.06] |
| rs114807731 | 3:60122415 | A | G | 0.022 | 2.98353×10^-5^ | 2.10734×10^-6^ | 1.99 [1.37-2.9] | 1.29 [1.09-1.53] | 1.68 [1.36-2.07] | 0.84 [0.616-1.15] |

**Supp. Table 3:** Top variants from the cardiometabolic (T2D and BMI) analysis (joint and interaction tests) along with stratified genetic odds ratios.

| **RSID** | **location** | **Effect Allele** | **Non-**  **Effect Allele** | **Effect Allele**  **Freq** | **Interaction p-value** | **Joint p-value** | **OR combined** | **OR (No T2D)** | **OR (T2D)** | **OR (No obesity)** | **OR (obesity)** |
| --- | --- | --- | --- | --- | --- | --- | --- | --- | --- | --- | --- |
| rs148793499 | 18:58314588 | C | T | 0.0102753 | 8.38293×10^-6^ | 1.32208×10^-8^ | 1.61 [1.27-2.03] | 1.56 [1.21-2.01] | 2.01 [1.03-3.93] | 1.18 [0.835-1.67] | 2.36 [1.7-3.27] |
| rs11115199 | 12:82510665 | T | G | 0.0204524 | 1.37218×10^-8^ | 4.85379×10^-8^ | 0.91 [0.736-1.12] | 0.72 [0.561-0.924] | 2.6 [1.73-3.91] | 0.96 [0.742-1.25] | 0.85 [0.592-1.22] |
| rs182113773 | 7:20239837 | A | C | 0.0147751 | 0.0529514 | 2.71316×10^-8^ | 1.73 [1.42-2.1] | 1.62 [1.31-2.01] | 2.6 [1.57-4.3] | 1.63 [1.26-2.11] | 1.94 [1.43-2.64] |
| rs2268616 | 14:75419444 | G | A | 0.0179483 | 0.26394 | 3.87431×10^-8^ | 1.64 [1.38-1.94] | 1.6 [1.33-1.92] | 2.01 [1.22-3.32] | 1.82 [1.48-2.24] | 1.38 [1.01-1.88] |
| rs6035512 | 20:19990781 | A | C | 0.0247661 | 0.000114499 | 8.88795×10^-8^ | 1.41 [1.2-1.66] | 1.45 [1.22-1.72] | 1.15 [0.648-2.03] | 1.21 [0.97-1.52] | 1.74 [1.36-2.22] |
| rs141850011 | 13:61925813 | C | T | 0.0103374 | 1.20059×10^-7^ | 2.72447×10^-7^ | 1.17 [0.89-1.53] | 0.88 [0.639-1.22] | 3.67 [2.22-6.05] | 0.97 [0.669-1.41] | 1.53 [1.03-2.26] |
| rs2727176 | 15:100570438 | G | A | 0.0102964 | 1.48748×10^-7^ | 2.84132×10^-7^ | 1.21 [0.931-1.57] | 0.94 [0.686-1.28] | 3.79 [2.3-6.24] | 0.97 [0.669-1.4] | 1.56 [1.07-2.27] |
| rs6747163 | 2:60777763 | G | C | 0.178678 | 2.6503×10^-7^ | 4.39×10^-7^ | 1.05 [0.977-1.14] | 0.99 [0.912-1.07] | 1.72 [1.39-2.12] | 1.11 [1.01-1.22] | 0.97 [0.858-1.11] |
| rs5747202 | 22:17969179 | G | T | 0.0374612 | 6.03274×10^-6^ | 2.70082×10^-7^ | 1.31 [1.1-1.56] | 1.15 [0.945-1.4] | 2.92 [1.93-4.42] | 1.26 [1-1.58] | 1.4 [1.05-1.85] |
| rs28864791 | 13:91502951 | C | T | 0.306404 | 5.06161×10^-5^ | 2.78527×10^-7^ | 0.89 [0.835-0.949] | 0.85 [0.792-0.908] | 1.32 [1.09-1.59] | 0.87 [0.805-0.948] | 0.91 [0.82-1.01] |
| rs144282550 | 4:74917179 | G | A | 0.0117938 | 0.00319523 | 2.91809×10^-7^ | 1.65 [1.32-2.06] | 1.51 [1.19-1.92] | 2.97 [1.72-5.12] | 1.65 [1.25-2.18] | 1.7 [1.19-2.43] |
| rs180972330 | 21:40008094 | G | A | 0.0100758 | 3.54×10^-7^ | 1.51×10^-6^ | 1.03 [0.774-1.38] | 0.79 [0.555-1.11] | 3.72 [2.18-6.35] | 0.71 [0.454-1.1] | 1.64 [1.11-2.41] |
| rs113083888 | 15:91197671 | T | C | 0.0484105 | 3.65×10^-7^ | 1.03×10^-6^ | 1.06 [0.933-1.21] | 1.12 [0.977-1.28] | 0.6 [0.354-1.02] | 0.95 [0.793-1.13] | 1.27 [1.04-1.55] |
| rs796925608 | 13:68788267 | ACTTTGAAAATAAC | A | 0.897704 | 0.100633 | 3.93×10^-7^ | 0.76 [0.681-0.839] | 0.77 [0.689-0.86] | 0.66 [0.487-0.902] | 0.75 [0.653-0.852] | 0.77 [0.647-0.912] |
| rs345361 | 4:86777776 | G | A | 0.969044 | 0.00084641 | 4.93×10^-7^ | 0.73 [0.63-0.84] | 0.79 [0.672-0.922] | 0.43 [0.296-0.611] | 0.75 [0.624-0.905] | 0.71 [0.561-0.896] |

Obesity was defined as BMI ≥ 30 for stratification.

**Supp. Table 4:** Top variants from the multiple deprivation index analysis (joint and interaction tests) along with stratified genetic odds ratios.

| **RSID** | **location** | **Effect Allele** | **Non-Effect Allele** | **Effect Allele Frequency** | **Interaction p-value** | **Joint p-value** | **OR interaction** | **OR combined** | **OR (Low MDI)** | **OR (High MDI)** |
| --- | --- | --- | --- | --- | --- | --- | --- | --- | --- | --- |
| rs147319526 | 1:231731679 | G | A | 0.012486 | 5.93×10^-8^ | 4.1588×10^-7^ | 3.13 [1.41-6.91] | 0.98 [0.746-1.3] | 0.44 [0.212-0.893] | 1.35 [0.961-1.89] |
| rs76612061 | 15:49972935 | A | G | 0.026409 | 6.25×10^-8^ | 2.1732×10^-7^ | 1.95 [1.2-3.19] | 0.85 [0.697-1.03] | 0.56 [0.363-0.853] | 1.08 [0.848-1.37] |
| rs6966810 | 7:20141514 | A | G | 0.929889 | 0.00946 | 1.6439×10^-7^ | 0.88 [0.694-1.1] | 0.8 [0.724-0.889] | 0.84 [0.692-1.01] | 0.72 [0.632-0.827] |
| rs73164732 | 7:144481768 | A | G | 0.011531 | 3.07×10^-7^ | 5.914×10^-7^ | 2.53 [1.26-5.09] | 1.17 [0.91-1.51] | 0.62 [0.332-1.17] | 1.6 [1.18-2.17] |
| rs2268616 | 14:75419444 | G | A | 0.018136 | 0.164618 | 3.3998×10^-7^ | 0.8 [0.548-1.17] | 1.64 [1.38-1.94] | 1.89 [1.41-2.53] | 1.5 [1.17-1.92] |
| rs1646599 | 18:55236161 | A | G | 0.01178 | 5.4×10^-7^ | 6.8523×10^-7^ | 0.96 [0.841-1.1] | 1.05 [0.986-1.11] | 1.06 [0.948-1.17] | 1.02 [0.938-1.1] |
| rs139020188 | 4:35620323 | T | A | 0.010373 | 6.03×10^-7^ | 1.6804×10^-6^ | 2.55 [1.19-5.47] | 1.08 [0.81-1.45] | 0.62 [0.312-1.23] | 1.56 [1.11-2.18] |
| rs148817892 | 16:5934373 | G | C | 0.011508 | 6.27546×10^-7^ | 2.51681×10^-6^ | 4.25 [1.85-9.79] | 1.1 [0.85-1.43] | 0.38 [0.176-0.835] | 1.62 [1.19-2.18] |
| rs72752741 | 9:89168171 | T | C | 0.014311 | 0.000798 | 7.014×10^-7^ | 1.08 [0.685-1.7] | 1.5 [1.23-1.84] | 1.45 [0.996-2.11] | 1.63 [1.25-2.13] |
| rs117488928 | 15:49563001 | C | A | 0.036988 | 9.93601×10^-7^ | 5.7925×10^-6^ | 1.79 [1.22-2.62] | 0.96 [0.822-1.12] | 0.69 [0.498-0.962] | 1.23 [1.02-1.5] |
| rs147349057 | 6:1059829 | TA | T | 0.015148 | 2.01×10^-6^ | 1.2453×10^-6^ | 2.43 [1.34-4.42] | 1.23 [0.98-1.54] | 0.69 [0.405-1.17] | 1.68 [1.28-2.22] |
| rs139833210 | 7:10216456 | A | T | 0.012896 | 0.001165 | 1.2661×10^-6^ | 0.6 [0.377-0.954] | 1.41 [1.13-1.77] | 2.17 [1.55-3.03] | 1.29 [0.933-1.78] |
| rs192911167 | 2:89058008 | G | C | 0.010853 | 1.57004×10^-6^ | 6.31539×10^-6^ | 2.55 [1.21-5.38] | 1.22 [0.932-1.59] | 0.59 [0.301-1.14] | 1.5 [1.08-2.08] |
| rs113541905 | 15:59709733 | T | C | 0.043553 | 1.72579×10^-6^ | 6.31187×10^-6^ | 1.57 [1.12-2.18] | 1.04 [0.903-1.19] | 0.8 [0.602-1.06] | 1.25 [1.04-1.48] |
| rs11945368 | 4:37555712 | A | C | 0.17519 | 0.0293873 | 1.9631×10^-6^ | 1.12 [0.948-1.33] | 1.18 [1.1-1.28] | 1.12 [0.979-1.29] | 1.26 [1.14-1.39] |

Low and high MDI were defined as being below or above the median value, respectively.
